# A simple algorithm based on initial Ct values predicts the duration to SARS-CoV-2 negativity and allows more efficient test-to-release and return-to-work schedules

**DOI:** 10.1101/2022.04.04.22273384

**Authors:** Olympia E. Anastasiou, Vu Thuy Khanh Le-Trilling, Mirko Trilling

## Abstract

Especially during global pandemics but also in the context of epidemic waves, the capacity for diagnostic qRT-PCRs rapidly becomes a limiting factor. Furthermore, excessive testing incurs high costs and can result in an overstrained work force in diagnostics departments. Obviously, people aim to shorten their isolation periods, hospitals need to discharge convalescent people, and re-employ staff members after infection. The aim of the study was to optimize retesting regimens for test-to-release from isolation and return-to-work applications. For this purpose, we investigated the association between Ct values at the first diagnosis of SARS-CoV-2 infection and the period until test negativity was reached, or at least until the Ct value exceeded 30, which is considered to indicate the transition to a non-infectious state. We included results from the testing of respiratory material samples for the detection of SARS-CoV-2 RNA, tested from 01 March 2020 to 31 January 2022.

Lower initial Ct values were associated with longer periods of SARS-CoV-2 RNA positivity. Starting with Ct values of <20, 20-25, 25-30, 30-35, and >35, it took median intervals of 20 (interval: 14-25), 16 (interval: 10-21), 12 (interval: 7-16), 7 (interval: 5-14), and 5 (interval: 2-7) days, respectively, until the person tested negative. Accordingly, a Ct threshold of 30 was surpassed after 13 (interval: 8-19), 9 (interval: 6-14), 7 (interval: 6-11), 6 (interval: 4-10), and 3 (interval: 1-6) days, respectively, in individuals with aforementioned start Ct values. Furthermore, the time to negativity was longer for adults versus children, wild-type SARS-CoV-2 variant versus other variants of concern, and in patients who were treated in the intensive care units.

Based on these data, we propose an adjusted retesting strategy according to the initial Ct value in order to optimize available PCR resources.

## Introduction

More than 5 billion COVID-19 tests have been performed worldwide since the beginning of the pandemic ^1^. Frequent testing and the isolation of infected individuals have been cornerstones of the global strategy to combat the COVID-19 pandemic. Rules for ending the isolation of infected individuals varied over time in different settings.

The European Centre for Disease Prevention and Control (ECDC) presents a differentiated approach to ending the isolation depending on patient specific characteristics (vaccination, immune status, severity of symptoms, closed vulnerable population settings) but also the pressure on healthcare systems and the society. Rapid antigen detection test is seen as an equivalent to real-time-PCR. Quarantine time varies from 10 to 20 days, but may be abridged though two negative antigen or PCR tests with a minimum 24-hour interval in most cases. A Ct value of over 30 can be used as cut-off of low likelihood of transmissibility in cases of prolonged PCR positivity ^2^.

A similar strategy is recommended by the German Robert-Koch-Institute, where a distinction is made between the general population including healthcare personnel, and patients. Release from isolation for the general population is possible after 10 days without any testing and with a negative antigen or PCR test or a PCR Ct value over 30 ^3^. For patients, the strategy differs according to their symptoms. For asymptomatic or mildly symptomatic individuals, release from isolation is possible after 14 days with a negative antigen test. PCR testing is reserved for patients with severe disease, where release from isolation is possible 14 days with a negative PCR result or a positive PCR, which is below a predefined level corresponding to 10^6^ copies/ml ^4^. An exception to these rules are immunosuppressed individuals and residents of age care facilities, where a decision is made from case to case ^5^.

Frequent testing in a hospital setting can to discharge convalescent individuals in a timely manner, which increases the number of available beds, and help to rapidly reemploy staff members after infection. On the other hand, it incurs high costs for both reagents and personnel. Bearing in mind that laboratory and personnel capacities are finite, increasing testing for SARS-CoV-2 may (and did for a time) limit molecular diagnostics for other infectious agents due to missing reagents or personnel constraints. The aim of this study was to investigate the relationship between initial virus loads and the duration of the infection. Our data show that the initial Ct value is highly predictive for the period until SARS-CoV-2 negativity or at least non-infectiousness will be reached. This information enables more efficient retesting regimens during times in which testing capacities are limited, and saves resources.

## Materials and Methods

We included results from the testing of respiratory material samples for the detection of SARS-CoV-2 RNA, tested from 01 March 2020 to 31 January 2022 at the Institute for Virology, University Hospital Essen, Germany.

Detection of SARS-CoV-2 RNA was performed using the RealStar SARS-CoV-2 RT-PCR kit (Altona Diagnostics, Hamburg, Germany), Alinity m SARS-CoV-2 Assay (Abbott, Wiesbaden, Germany), Abbott RealTime SARS-CoV-2 assay (Abbott, Wiesbaden, Germany), Xpert Xpress SARS-CoV-2 assay (Cepheid, Krefeld, Germany) and Xpert Xpress SARS-CoV-2/Flu/RSV (Cepheid, Krefeld, Germany). For the Ct values, the values for the envelope (E) gene were used for samples tested with the RealStar^®^ SARS-CoV-2 RT-PCR kit and the Xpert Xpress SARS-CoV-2 assay. For the results generated with the Abbott RealTime SARS-CoV-2 assay, 10 was added to the Ct value, since the first 10 PCR cycles in this assay are unread ^6^.

The date of diagnosis was defined as the date of the first positive SARS-CoV-2 sample in the database. Date of negativity was defined as date of the first negative SARS-CoV-2 sample after the last positive sample. Date of Ct > 30 was defined as the date of the first negative sample or positive sample with Ct > 30 (whichever came first) after first diagnosis. Excluded were negative samples where the time period between the last positive sample and first negative exceeded 7 days and also positive or negative samples where the time period between the first diagnosis and last positive sample date exceeded 90 days to exclude reinfections.

Initially, 109,264 individuals were included in the analysis. For 947 of them, we had a negative result after SARS-CoV-2 and 1,312 had a negative result or positive result with a Ct value over 30 meeting aforementioned criteria. Furthermore, the time from March 2020 to January 2022 was stratified into 4 periods according to the SARS-CoV-2 variant found in the majority of samples in each calendar week. The tested samples were evaluated with melting curve analysis for the SNPs N501Y, L452R, E484K, and/or S137L (TIB MOL BIOL, Berlin Germany), partial sequencing of the spike protein (S) gene (Sanger Sequencing) and/or whole genome sequencing (Next Generation Sequencing, Illumina MiSeq). The first time period spanned from the beginning of the pandemic until week 8 of 2021, when the wild type variant was the most prevalent virus variant, the second period ranged from week 9 of 2021 to week 27 of 2021, during which the alpha variant predominated, the third period from week 28 of 2021 to week 52 of 2021, when the delta variant circulated, and the fourth period from week 1 of 2022 to end of January 2022, when the omicron variant was most frequent.

The ethics committee of the medical faculty of the University of Duisburg-Essen approved the analysis of data for the improvement of diagnostic procedures (20-9512-BO). Statistical analyses were performed using SPSS software (v23, SPSS Inc., Chicago, IL, USA) and GraphPad Prism 6.0 (GraphPad, CA, USA). Normal distribution was evaluated using the Shapiro-Wilks test. Comparisons between groups were performed using Mann-Whitney-U or Kruskal-Wallis (adjusted for multiple comparisons) tests as applicable. Two-tailed *p* values less than 0.05 were considered statistically significant.

## Results

### Time to negativity is longer for adults vs children, wild-type SARS-CoV-2 variant vs other variants and patients in the intensive care unit

Overall, the median time to negativity in days was 11 with an interquartile range (IQR) of 4 to 21. There was no difference between men and women, while children tended to have a shorter time to negativity compared to adults [4 (1-14) vs 12 (4-21), p<0.001]. After stratifying our patients according to their age, children younger than 10 years of age had a significantly shorter time to negativity compared to adults aged from 50-59, 60-69, 70-79, 80-89 years. People aged 10-19 years had a shorter time to negativity compared to the 80-89 years old group. Patients, who had been treated at any point in an intensive care unit (ICU) had a longer time to negativity compared to those who did not (Table 1, Figure 1A). A similar pattern was evident, when focusing on the period after which the Ct surpassed 30 (Figure 1B).

**Table 1:** Time to negativity or Ct>30 in our cohort.

| Group | Time to negativity (days) |  |  | Time to Ct>30 (days) |  |  |
| --- | --- | --- | --- | --- | --- | --- |
|  | n | Median (IQR) | p | n | Median (IQR) | P |
| All | 947 | 11 (4-21) | - | 1317 | 7 (3-13) |  |
| **Sex** |  |  |  |  |  |  |
| male | 464 | 12 (4-23) |  | 653 | 7 (3-13) |  |
| female | 480 | 10 (3-19) | 0.081 | 653 | 7 (3-13) | 0.682 |
| **Adults vs Children** |  |  |  |  |  |  |
| adults | 897 | 12 (4-21) |  | 1241 | 7 (4-13) |  |
| children | 50 | 4 (1-14) | <0.001 | 71 | 5 (1.5-8.5) | 0.002 |
| **Age** |  |  |  |  |  |  |
| 0-9 years | 20 | 2 (1-8) | - | 32 | 4.5 (1-13) | NS |
| 10-19 years | 42 | 5 (2-16) | vs 80-89<br>0.049 | 53 | 6 (2-9) | NS |
| 20-29 years | 109 | 7 (2-18) | NS | 143 | 6 (2-10) | vs 60-69<br>0.029 |
| 30-39 years | 116 | 7,5 (3-16.5) | NS | 152 | 7 (3-10) | NS |
| 40-49 years | 123 | 8 (3-20) | NS | 167 | 7 (3-12) | NS |
| 50-59 years | 164 | 12 (4-23.5) | vs 0-9 years<br>0.04 | 247 | 8 (4-13) | NS |
| 60-69 years | 154 | 12 (6-25) | vs 0-9 years<br>0.012 | 220 | 8 (5-13) | - |
| 70-79 years | 110 | 14 (5-55) | vs 0-9 years<br>0.031 | 149 | 7 (4-13) | NS |
| 80-89 years | 92 | 14,5 (5-27) | vs 0-9 years<br>0.006 | 129 | 7 (4-14) | NS |
| > 90 years | 17 | 18 (4-22) | NS | 20 | 7 (4-12) | NS |
| **Time period with** |  |  |  |  |  |  |
| Majority variant wild type | 530 | 13 (6-24) | - | 759 | 8 (5-13) | - |
| Majority variant alpha | 167 | 7 (2.5-17.5) | vs wt<br><0.001 | 227 | 6 (2-9.5) | vs wt<br><0.001 |
| Majority variant delta | 178 | 6.5 (1-22) | vs wt<br><0.001 | 249 | 7 (1-12) | vs wt<br><0.001 |
| Majority variant omicron | 72 | 7 (2-14) | vs wt<br><0.001 | 77 | 7 (2-8) | vs wt<br><0.001 |
| **Intensive Care Unit (ICU)** |  |  |  |  |  |  |
| patients in ICU | 296 | 14 (6-26) |  | 435 | 8 (4-14) |  |
| other patients | 651 | 8 (3-19) | <0.001 | 877 | 7 (3-12) | 0.001 |

IQR = interquartile range; NS = not significant. Where multiple comparisons were performed, only pairwise comparisons that remained statistically significant after correction for multiple comparisons are shown in the table.

**Figure 1:**
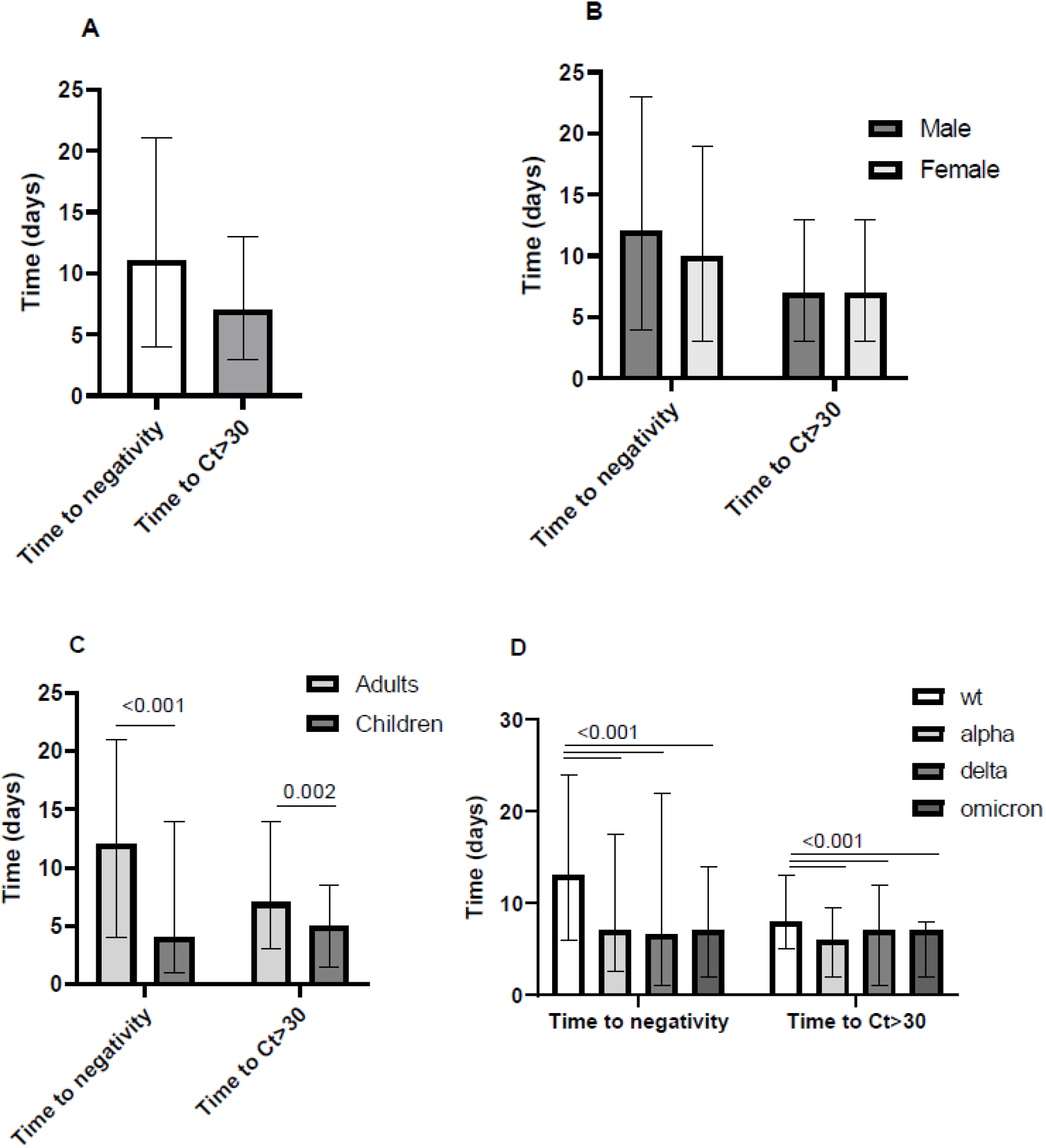
Time to negativity in days or to a positive result with a Ct>30 since the first SARS-CoV-2 positive result in the whole cohort (A), in male versus female subjects (B) in adults versus children (C) and in time periods, when wild type (wt), alpha, delta, and omicron SARS-CoV-2 variants were dominant (D).

### Lower Ct values are associated with longer time to SARS-CoV-2 RNA undetectability

Next, we focused on the period to the first negative result or the first result with Ct>30 after diagnosis in regard to the Ct value of previous samples. Lower Ct values were associated with longer time to the first negative or low positive result (Ct >30) (Table 2). All comparisons concerning time to negativity between the different Ct value groups showed a statistical significance with the exception of groups Ct<20 versus Ct 20-25. All comparisons concerning time to a Ct over 30 after diagnosis between the different Ct value groups showed a statistical significance (Supplement, Table S1).

**Table 2:** Time to the first negative result and result with a Ct>30 according to the stratified Ct values of previous SARS-CoV-2 positive samples.

| Group | Time to first negative result (days) |  | Time to Ct>30 (days) |  |
| --- | --- | --- | --- | --- |
|  | n | Median (IQR) | n | Median (IQR) |
| Ct <20 | 154 | 19.5 (14-25) | 279 | 13 (8-19) |
| Ct 20-24.99 | 195 | 16 (10-21) | 327 | 9 (6-14) |
| Ct 25-29.99 | 240 | 11.5 (7-16) | 346 | 7 (6-11) |
| Ct 30-34.99 | 341 | 7 (5-14) | 207 | 6 (4-10) |
| Ct>35 | 405 | 5 (2-7) | 217 | 3 (1-6) |

IQR= interquartile range.

This effect was more pronounced, when looking at samples from the first phase of the pandemic, when the dominant SARS-CoV-2 variant was wild type compared to the next phases (Table 3, Figure 2). There was no significant difference of the time to the first negative result and time to Ct>30 according to the stratified Ct values of previous SARS-CoV-2 positive samples in men vs women or patients treated in an intensive care unit vs other patients (data not shown). An age-based stratification and comparison was not possible due to the scarcity of data. All comparisons concerning time to negativity between the different Ct value groups showed a statistical significance with the exception of groups Ct<20 vs Ct 20-25 and Ct 25-30 versus Ct 30-35 for the wt time period and groups Ct<20 versus Ct 20-25 and groups Ct<20 and Ct 25-30 for the non wt period. All comparisons concerning time to a Ct value over 30 after diagnosis between the different Ct value groups showed a statistical significance with the exception of groups Ct 25-30 versus Ct 30-35 for the wt time period, groups Ct 20-25 versus Ct 25-30 and groups Ct<20 versus Ct 20-25 for the non wt time period (Supplement, Table S1).

**Table 3:** Time to first negative result and time to Ct>30 according to the stratified Ct values of previous SARS-CoV-2 positive samples, at different time-periods of the pandemic.

| **Time period with** | **Majority variant wild type** |  | **Majority variant non wild type** |  |  |
| --- | --- | --- | --- | --- | --- |
| Group | n | Median (IQR) | n | Median (IQR) | p |
| **Time to the first negative result (days)** |  |  |  |  |  |
| Ct <20 | 107 | 21 (14.5-25.5) | 47 | 15 (12-22.5) | **0.035** |
| Ct 20-24.99 | 131 | 18 (10.5-23.5) | 64 | 14 (9-19.5) | **0.021** |
| Ct 25-29.99 | 146 | 11 (7-16) | 94 | 12 (7-15) | 0.687 |
| Ct 30-34.99 | 213 | 7 (6-14) | 128 | 6 (4-14) | 0.067 |
| Ct>35 | 216 | 6 (4-9) | 189 | 4 (2-7) | **0.001** |
| **Time to Ct>30 (days)** |  |  |  |  |  |
| Ct <20 | 196 | 13 (10-19) | 83 | 11 (7-18) | **0.007** |
| Ct 20-24.99 | 214 | 9.5 (4-15) | 113 | 8 (5-13) | 0.065 |
| Ct 25-29.99 | 214 | 7 (6-11) | 132 | 7 (6-11.5) | 0.726 |
| Ct 30-34.99 | 127 | 7 (5-10) | 80 | 6 (2.5-9.5) | **0.039** |
| Ct>35 | 102 | 4 (1-.6) | 115 | 2 (1-6) | **0.02** |

IQR= interquartile range.

**Figure 2:**
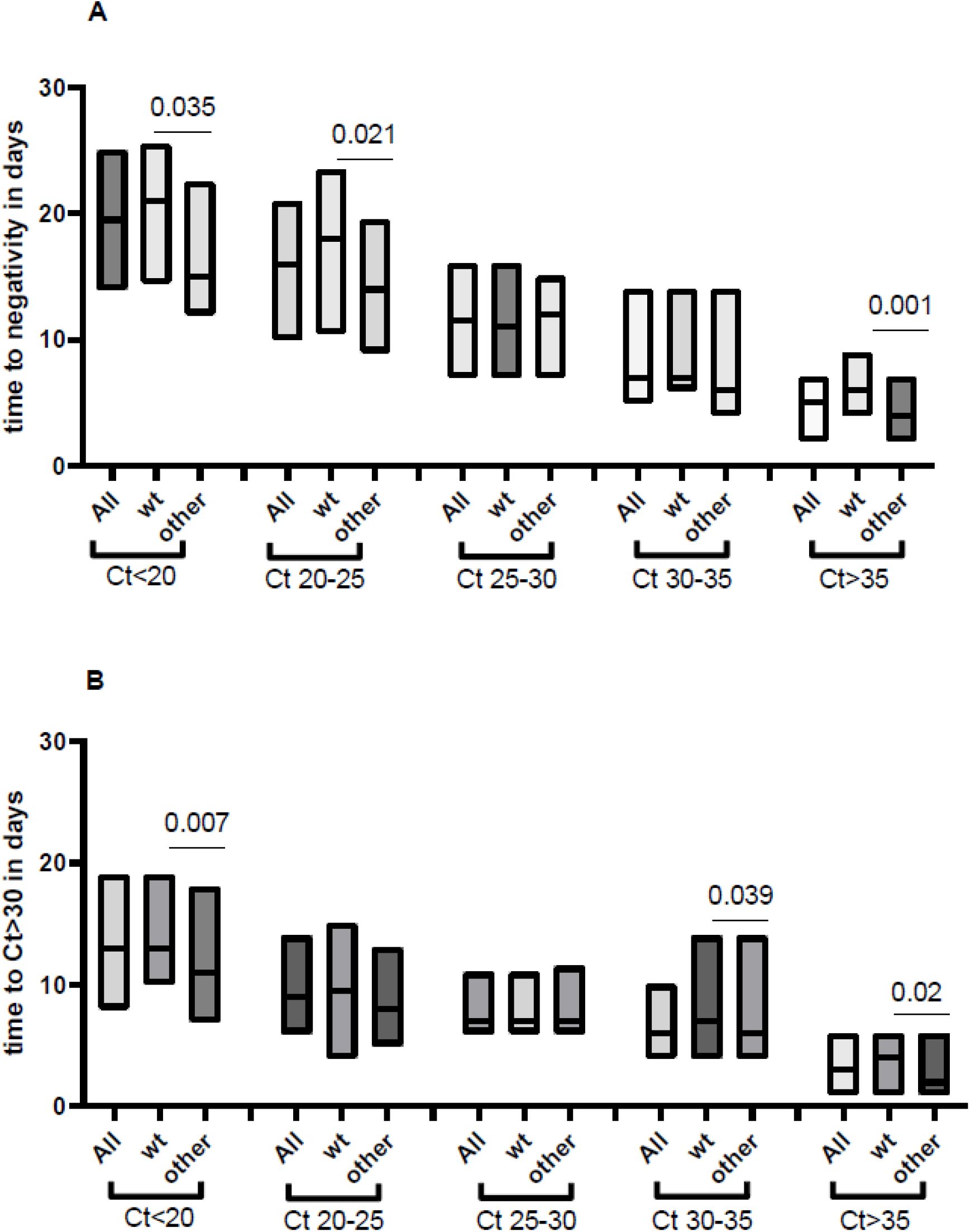
Time to negativity in days (A) or to a positive result with a Ct>30 (B) in conjunction with the Ct value of previous samples. Wild type (wt) vs other refer to periods, when the respective SARS-CoV-2 variants were dominant. In (A), all comparisons concerning time to negativity between the different Ct value groups showed a statistical significance with the exception of groups Ct<20 versus Ct 20-25 for the whole cohort as well as the two different time periods, groups Ct 25-30 versus Ct 30-35 for the wt time period, groups Ct 20-25 versus Ct 25-30 and groups Ct <20 versus Ct 25-30 for the non wt period. In (B), all comparisons concerning time to a Ct value over 30 after diagnosis between the different Ct value groups showed a statistical significance with the exception of groups Ct 25-30 versus Ct 30-35 for the wt time period, groups Ct 20-25 versus Ct 25-30 and groups Ct <20 versus Ct 20-25 for the non wt time period.

## Discussion

Our data indicate that viral shedding was more prolonged in adults as compared to children. This is in accordance with previous studies, which indicate that age is positively correlated with the duration of viral shedding in SARS-CoV-2 infection ^7,8^. We also found that patients who needed treatment in the intensive care unit had longer shedding periods compared to other patients, in concordance with previous data ^9^. Cases from the first phase of the pandemic, when the dominant SARS-CoV-2 variant was wild type, had a longer time to negativity compared to cases from other time periods, when the alpha, delta or omicron variant were dominant. Interestingly time to negativity or time to reaching a lower viral load (Ct>30) were very similar, when comparing data from the phases in the pandemic when alpha, delta or omicron were dominant. Previous data comparing the time to negative PCR in patients infected with omicron versus delta variant indicated also that there is no significant difference between the two groups ^10^.

Lower Ct values in tested samples were associated with longer time to SARS-CoV-2 RNA undetectability. This effect was more pronounced in the first period of the pandemic with wild type as majority variant. Focusing on the following period of the pandemic, when alpha, delta and omicron variants were dominant, only a quarter of cases with a Ct values of less than 20 would have a PCR reversion in a 12 days time, most of the patients would need more than 2 weeks. Also, retesting patients with Ct values of 20-25 in a week or 10 days does not seem efficient, since only 25% of them become negative after 9 days. On the contrary, retesting in a week seems to be make sense for patients with Ct values greater than 30. If one uses the Ct >30 limit as the criterion for lifting some restrictions, then retesting in a week would make sense in cases with a Ct value greater than 25, but it would still be too soon for patients with lower Ct values.

This analysis has some limitations. We have no data on patient symptoms (including severity and time of onset) and some of the cases may have been diagnosed before as SARS-CoV-2 positive, prior to admittance. However, our data provide a valuable insight on the dynamic of viral shedding of SARS-CoV-2.

To date, more than 5 billion COVID-19 tests have been performed worldwide since the beginning of the pandemic ^1^. The gold standard for SARS-CoV-2 diagnostics is the real-time PCR ^11^. It requires specialized equipment and personnel, is expensive and due to rapid increase in its use shortages in reagents have been observed. In an effort to reduce PCR testing and thus preserve resources, rapid SARS-CoV-2 tests have been recommended as an alternative in many but not all cases ^2,5^. Notwithstanding the concerted effort to reduce PCR testing, it has been our experience that testing in ours and other hospitals is more rigorous and less uniform in different departments than official national recommendations (see Introduction). A negative PCR result is more often than not necessary to release a patient from isolation, health care personnel can resume their duties with a negative or low positive (Ct>30) PCR result. Adapting the testing PCR strategy according to previous Ct values could be a way to save laboratory and personnel resources. It could be also used to manage hospital resources (personnel, bed capacity) more efficiently.

## Data Availability

Additional aggregated data produced in the present study are available upon reasonable request to the authors

