## Supplement for "A simple algorithm based on initial Ct values predicts the duration to SARS-CoV-2 negativity and allows more efficient test-to-release and return-to-work schedules"

Table S1: P-values of the pairwise comparisons of time to the first negative result and result with a Ct>30 according to the stratified Ct values of previous SARS-CoV-2 positive samples, overall and at different timepoints (wild type vs non wild type period).

| Time to the first negative result, whole cohort | | | | | |
| --- | --- | --- | --- | --- | --- |
| Ct values | <20 | 20-25 | 25-30 | 30-35 | >35 |
| <20 | - | 0.053 | **<0.001** | **<0.001** | **<0.001** |
| 20-25 | 0.053 | - | **0.001** | **<0.001** | **<0.001** |
| 25-30 | **<0.001** | **0.001** | - | **0.002** | **<0.001** |
| 30-35 | **<0.001** | **<0.001** | **0.002** | - | **<0.001** |
| >35 | **<0.001** | **<0.001** | **<0.001** | **<0.001** | - |
| Time to the first negative result, wt time period | | | | | |
| Ct values | <20 | 20-25 | 25-30 | 30-35 | >35 |
| <20 | - | 0.210 | **<0.001** | **<0.001** | **<0.001** |
| 20-25 | 0.210 | - | **<0.001** | **<0.001** | **<0.001** |
| 25-30 | **<0.001** | **<0.001** | - | 0.318 | **<0.001** |
| 30-35 | **<0.001** | **<0.001** | 0.318 | - | **<0.001** |
| >35 | **<0.001** | **<0.001** | **<0.001** | **<0.001** | - |
| Time to the first negative result, non wt time period | | | | | |
| Ct values | <20 | 20-25 | 25-30 | 30-35 | >35 |
| <20 | - | 1 | 0.133 | **<0.001** | **<0.001** |
| 20-25 | 1 | - | 1 | **0.001** | **<0.001** |
| 25-30 | 0.133 | 1 | - | **0.008** | **<0.001** |
| 30-35 | **<0.001** | **0.001** | **0.008** | - | **<0.001** |
| >35 | **<0.001** | **<0.001** | **<0.001** | **<0.001** | - |
| Time to the positive result with Ct>30 after diagnosis, whole cohort | | | | | |
| Ct values | <20 | 20-25 | 25-30 | 30-35 | >35 |
| <20 | - | **<0.001** | **<0.001** | **<0.001** | **<0.001** |
| 20-25 | **<0.001** | - | **0.019** | **<0.001** | **<0.001** |
| 25-30 | **<0.001** | **0.019** | - | **0.008** | **<0.001** |
| 30-35 | **<0.001** | **<0.001** | **0.008** | - | **<0.001** |
| >35 | **<0.001** | **<0.001** | **<0.001** | **<0.001** | - |
| Time to the positive result with Ct>30 after diagnosis, wt time period | | | | | |
| Ct values | <20 | 20-25 | 25-30 | 30-35 | >35 |
| <20 | - | **<0.001** | **<0.001** | **<0.001** | **<0.001** |
| 20-25 | **<0.001** | - | **0.006** | **<0.001** | **<0.001** |
| 25-30 | **<0.001** | **0.006** | - | 0.810 | **<0.001** |
| 30-35 | **<0.001** | **<0.001** | 0.810 | - | **<0.001** |
| >35 | **<0.001** | **<0.001** | **<0.001** | **<0.001** | - |
| Time to the positive result with Ct>30 after diagnosis, non wt time period | | | | | |
| Ct values | <20 | 20-25 | 25-30 | 30-35 | >35 |
| <20 | - | 0.065 | **0.008** | **<0.001** | **<0.001** |
| 20-25 | 0.065 | - | 1 | **0.002** | **<0.001** |
| 25-30 | **0.008** | 1 | - | **0.008** | **<0.001** |
| 30-35 | **<0.001** | **0.002** | **0.008** | - | **0.001** |
| >35 | **<0.001** | **<0.001** | **<0.001** | **0.001** | - |
